# Epidemiological and phylogenetic analyses of public SARS-CoV-2 data from Malawi

**DOI:** 10.1101/2024.06.28.24309607

**Authors:** Mwandida Kamba Afuleni, Roberto Cahuantzi, Katrina A. Lythgoe, Atupele Ngina Mulaga, Ian Hall, Olatunji Johnson, Thomas House

## Abstract

The novel Coronavirus SARS-CoV-2 was first identified in a person in Wuhan city, China in December 2019, and had spread to all continents in less than three months. While there were many similarities between the resulting COVID-19 pandemic in different regions and countries, there were also important differences, motivating systematic quantitative analysis of available data for as diverse a set of geographical locations as possible to drive generation of insights relevant for response to COVID-19 and other respiratory viral and pandemic threats. Malawi had its first COVID-19 case on 2 April 2020 and, like many countries in the Global South, had access to orders of magnitude less data than countries in the Global North to inform its response. Here, we present modelling analyses of SARS-CoV-2 epidemiology and phylogenetics in Malawi from 2 April 2020 to 19 October 2022. We carried out this analysis using open-source software tools and open data on cases, deaths, geography, demographics, and viral genomics. In particular, we used R to visualise the raw data and results, alongside Generalised Additive Models (GAMs), which were fit to case and mortality data to describe the incidence trends, growth rate and doubling time of SARS-CoV-2. IQTree, TreeTime and interactive Tree of Life were used to perform the phylogenetic analysis. This analysis reveals five major waves of COVID-19 in Malawi, associated with different lineages: (1) Early variants; (2) Beta; (3) Delta; (4) Omicron BA.1; (5) Other Omicron. Some sequences associated with the Alpha variant were present but these did not appear to drive a major wave as they did in some other countries. Case Fatality Ratios were higher for Delta, and lower for Omicron, than for earlier lineages. Phylogeny reveals separation of the tree into major lineages as would be expected, and early emergence of Omicron, as is consistent with proximity to the likely origin of this variant. Both variant prevalence and overall rates of cases and deaths were highly geographically heterogeneous. We argue that such analyses could have been and could in future be carried out in real time in Malawi and other countries in the Global South with similar computational and data resources.

**Author summary:** Malawi detected its first infection with SARS-CoV-2 at the start of April 2020, and like many other countries in the Global South did not have comparable volumes of data to Global North countries to inform its response to the COVID-19 pandemic. Here, we present quantitative analyses of the epidemiology and phylogenetics of SARS-CoV-2 in Malawi using open software and data that can be straightforwardly deployed in other countries and for other pathogens, under similar data availability. We observed five major COVID waves over a period from April 2020 to October 2022, each associated with different variants of SARS-CoV-2, as well as significant geographical heterogeneity. Waves were typically associated with early doubling times of between 7 and 4 days, with the second major wave driven by the Beta variant rather than the Alpha and Gamma variants observed in some other countries. Pylogenetic analysis revealed a temporal tree structure consistent with both major variant structure identified elsewhere, and known epidemiology of major variants.

## Introduction

Emerging and re-emerging infectious diseases such as Tuberculosis (TB), HIV/AIDS, Dengue Fever, Ebola, Influenza, SARS, and MERS are a public health concern worldwide. Infectious diseases still top the list of causes of death in the world [1]. Recently, the whole world was hit by a novel Coronavirus, severe acute respiratory syndrome coronavirus-2 (SARS-CoV-2) [2, 3], which causes a disease known as COVID-19 [4]. SARS-CoV-2 attacks lung epithelial cells in human beings, through angio-tensionconverting enzyme 2 (ACE2) receptor. A person suffering from COVID-19 shows symptoms such as dry cough, sore throat, breathing difficulties, high temperature, diarrhoea, headache, muscle pain, joint pain, fatigue, and loss of sense of smell and taste[5, 4, 6]. COVID-19 first emerged in Wuhan city, China in December 2019. After the first case was reported, the virus spread to all continents within less than three months, with Africa being the last continent to be hit [4][6]. The World Health Organisation (WHO) declared coronavirus a Public Health Emergency of International Concern (PHEIC) on 30 January 2020 [7]. A year later, more than one hundred million people from over 210 countries had been infected, while over two million people had lost their lives to COVID-19. In Africa, the first COVID-19 case was reported on 14 February 2020 in Egypt and spread across the continent within three months. Lesotho was the last state in Africa to be affected [8]. SARS-CoV-2 spread to all continents, risking many lives and lowering global life expectancy[9].

According to the WHO, SARS-CoV-2 variants have one of six main names: Alpha; Beta; Gamma; Delta; Omicron; and Non Variants of concern (non-VOC), which throughout this study is referred to as ‘Other’. Delta, Omicron and Other in particular have numerous sub-variants [10]. Omicron was first identified in South Africa in November 2021, then later spread more rapidly to other countries. The rapid growth of Omicron resulted from the occurrence of the mutations on the spike protein, which increases the chance of infection[10]. Although Omicron is highly transmissible, disease severity is lower than for Delta. Delta emerged before Omicron and was first identified in India in late 2020. Delta spread across the whole world and its disease severity was double as much as that of the other variants. Literature shows that the original variant of Omicron spread more rapidly than Delta[10]. Alpha was more problematic than the original virus, it first emerged in Great Britain in November 2020, later spread to all parts of the world within a short period and became dominant in the U.S.[10]. Delta’s disease severity is higher than that of Alpha. In late 2020 in South Africa, the Beta variant was discovered for the first time and it was believed to have many mutations, more severe and caused more hospitalisations and deaths than the other variants[10]. The Gamma variant originated in Brazil in November 2020 [11]. Like Alpha and Beta, Gamma attaches to human cells easily but is not as infectious as Alpha or Delta[12].

Malawi registered its first COVID-19 case on 2 April 2020 [13], two weeks after declaring a state of national disaster on March 20, 2020 [14][15]. Following the first confirmed case, the Malawi government put in place COVID-19 restrictions that included; banning international travel, prohibiting public events, closing schools at all levels, decongesting workplaces (where people were encouraged to work from home) and reducing the capacity of public transport [16]. In addition, the government recommended wearing face masks, frequent washing of hands and massive testing of everyone showing signs and symptoms of COVID-19. Furthermore, the state made sure that the public was aware of the pandemic, its symptoms and the preventive measures, by using various communication mediums and vernacular languages [16]. A report by the International Food Policy Research Institute (IFPRI) of June 2020, described Malawi’s response to the COVID-19 pandemic as quite satisfactory[17]. Ngwira *et al* (2021) in their study carried out from April to October 2020 found that the infection risk was higher among People living in the major cities of Malawi than those living in rural areas. The authors attributed their findings to higher population density in the cities than in the rural areas, implying that the higher the population density, the faster the spread of the virus [18].

This work is to our knowledge the first comprehensive analysis of the disease epidemiology and phylogenetics of COVID-19 in Malawi using public data. We identify lineages associated with the major waves of COVID-19 and the time of most recent common ancestor (tMRCA) of the variants. Geographical heterogeneity of SARS-CoV-2 in Malawi is revealed, which is one of the important factors to consider when combating an infection outbreak. The paper further describes trends of SARS-CoV-2 cases and deaths; contrasts the infectiousness of the variants; and quantifies the infection’s case fatality rate for each wave.

The paper has been arranged in a way that the next section describes the data as well as the methods applied to the analyses. After the methods section, results from the analyses are presented and later discussed in the section that follows. The last section concludes the paper by providing the implications of the findings.

## Materials and methods

### Population and study setting

This study used data from Malawi. Malawi is a landlocked country that is bordered by Mozambique to the East, South and Southwest, Tanzania to the north and northeast and Zambia to the West and Northwest. The country is located at latitudes between 9*^◦^* 22’ S and 17*^◦^* 03’ S and longitudes between 33*^◦^* 40’ E and 35*^◦^* 55’ E. Malawi has a total surface area of 118,484 km^2^ [19] and a total population size of approximately 18 million [20]. The country has three regions; North, Central and South; and 28 districts. According to Malawi’s 2018 population and housing census the capital city, Lilongwe, which lies in the central region has the highest population proportion of 9.3%. More than 80% of the people live in rural areas and 16% in the urban areas. Half of the Malawi population is 17 years old or below [20]. Malawi is categorised as a Low-Income country [21] with a fragile health system [22]. The health system, which has four levels; community, primary, secondary and tertiary, lacks both financial and human resources [23]. In a year before the 2018 household census across the country, there were 6.3 all-cause deaths and 32.8 births per 1,000 persons [20].

### Data sources

Publicly available datasets on cases, deaths and genome of SARS-CoV-2 were used in this study. This means that the analysis should be completely reproducible by all researchers with Internet access and a modern desktop or laptop personal computer.

### Cases and deaths

SARS-CoV-2 was first identified in Malawi on 2 April 2020. New confirmed cases continue to be reported to date, although the numbers are relatively small. In this study, we use data on daily new cases of SARS-CoV-2 beginning from 2 April 2020 to 19 October 2022, and new deaths beginning from 7 April 2020 to 19 October 2022. This was extracted from the Our World in Data (OWID) website [24], and corresponds to a study period of about two and a half years.

### Genomic data for SARS-CoV-2

COVID-19 sequence data for Malawi was accessed from GISAID (the Global Initiative on Sharing All Influenza Data) [25, 26, 27], an open database. The data comprised of 1, 436 DNA sequences that were collected from 5 April 2020 to 19 January 2023. We downloaded the FASTA file containing the sequences and other files in TSV and TAR formats containing metadata including collection and submission date, host, additional host, location, genome, lineage, clade, gender, patient age, sampling strategy and AA substitutions, among others.

### Geographical and demographic data

For mapping, Shape files containing the boundaries of sub-national regions in vector format were obtained from the Humanitarian Data Exchange [28], the total SARS-CoV-2 confirmed cases in districts were obtained from the the dashboard of the public health institute of Malawi (PHIM)[29]u and demographic data on population sizes for these regions were obtained from the Malawi National Statistical Office [20].

### Data analysis

#### Case fatality rate (CFR) and growth rate analysis

We quantify and compare the disease severity amongst the variants, by computing CFR with 95% CrI derived using Bayesian conjugate inference for each variant. The ratio of the CFR’s and the uncertainty in this ratio is computed using Monte Carlo approximation methods, and from this percentage differences in CFR between pairs of variants are calculated. We fitted case and death data using R statistical package [30], particularly Generalised Additive Models (GAMs) [31] with log link function. We used these to calculate growth rate and doubling time following the approach of [32, 33], noting that if *s*(*t*) is a smoothed estimate of the logarithm of a mean signal, then its time derivative 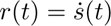 is an estimate of the instantaneous growth rate and *τ_D_* = log(2)*/r*(*t*) is an estimate of the instantaneous doubling time [34].

GAMs were applied particularly for their flexibility, reliability and validity when fit to non-linear and real-world data. Cases or deaths were a function of time (day and day-of-week) in the model. Day-of-week was considered because reported cases or deaths were seemingly low during weekends. Further details on how the model was fit to data as well as calculation of the instantaneous growth rate and doubling time of the infection are found in the Supplementary Material, S1.

#### National phylogenetic analysis

We used viral genomic data to identify SARS-CoV-2 variants, and TreeTime [35] to perform a timescaled phylogenetic analysis in order to describe the evolution of the virus including the time of the most recent common ancestor (tMRCA) for the variants. Before the analysis, Sequences in a FASTA file were categorised according to names that WHO and researchers agreed to give to specific COVID-19 lineages [36]. A total of 1,436 sequences were categorised into five variants as shown in a table in the Supplementary material S1. Multiple sequence alignment was performed using MAFFT [37] before a maximum likelihood tree was reconstructed using IQ-TREE [38], a stochastic algorithm that builds trees by maximum likelihood methods. The tree was later fit to a calendar and visualised using TreeTime version 0.82 [35].

#### Geographical analysis

We mapped confirmed cases, total population, and cases per capita using the R package cartography [39]. Sequence alignment and tree reconstruction were repeated for individual variants and were then annotated in interactive Tree Of Life (iTOL) [40] to show the regions the sequences were collected from to compare the distribution of the variants across the three regions of Malawi. We performed a *χ*^2^ test for the difference in proportions in R using the MASS package [41]. This analysis would be helpful in the future when similar risks arise since provides scientifically proven information to the authorities on whether or not the regions are equally affected, a useful factor to consider when allocating resources to the affected regions.

## Results

### Case Fatality Rate (CFR)

The overall CFR for the whole study period is 3.05% (95% credible interval (2.94, 3.16)%). Delta was more severe (CFR = 4.16%, CrI= (3.93, 4.40)) than the other variants while Omicron had the lowest CFR = 1.46%, CrI= (1.32, 1.61)%. Table 1 compares the CFR and their 95% credible intervals for the variants. The posterior distribution of CFR for the variants suggests the same, that Delta has the highest CFR and Omicron has the lowest. The full posterior distribution of CFR for the variants is shown in Supplementary Material S1, showing the CFR of Other and Beta overlapping. Further analysis of the ratios of CFRs, transformed to percentage changes, is shown in Figure 1.

**Table 1:** Case Fatality Rate for SARS-Cov-2 variants.

| Variant | Deaths | Cases | CFR (%) | 95% CrI |
| --- | --- | --- | --- | --- |
| Other | 185 | 6,043 | 3.07 | (2.66, 3.53) |
| Beta | 957 | 27,932 | 3.43 | (3.22, 3.65) |
| Delta | 1,160 | 27,871 | 4.16 | (3.93, 4.40) |
| Omicron | 381 | 26,218 | 1.46 | (1.32, 1.61) |

**Figure 1:**
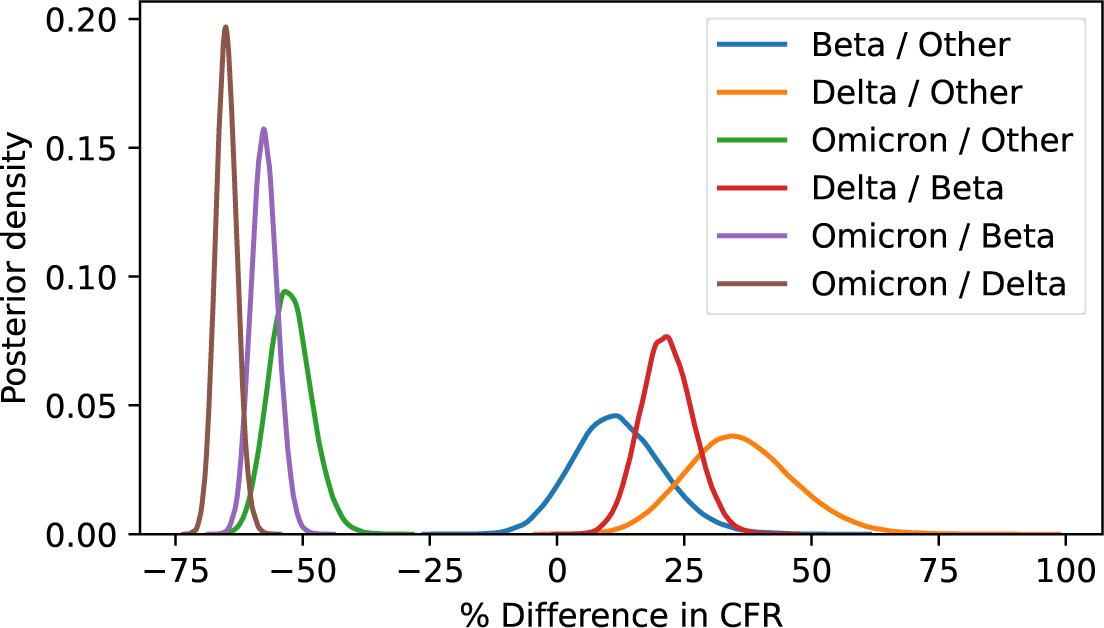
Case Fatality Ratio Differences. Posterior densities of the ratios of different strain CFRs, showing high credibility of increase for Delta and decrease for Omicron compared to everything else, but only weak evidence for a difference between Beta and ‘other’ strains.

### Trends of COVID-19 Reported Cases and Deaths in Malawi

#### COVID-19 reported cases modelling

As of 19 October 2022, Malawi had a total of 88, 064 confirmed cases of SARS-CoV-2. [24] Fig 2A shows the number of daily cases from 2 April 2020 to 19 October 2022. Five peaks are clearly visible in the plot, representing the five waves however, the last peak is very short. On average, there were 95 new reported cases every day, with 1, 316 as the highest observed number of cases and 0 as the lowest. The variance was 41, 200 and 95% of the values for daily new cases were below 547. The colours in Fig 2A, each represents a different wave dominated by a COVID-19 variant, which implies that one variant spanned over the last two waves. Dates at which waves started or new colour began were determined by the use of linear discriminant analysis (LDA). In that regard, the Beta wave started on December 3, 2020, the Delta wave on April 21, 2021 and Omicron on November 16, 2021. According to genome data, Malawi’s first variant was non-VOC, which emerged before Alpha (Other), second was Beta followed by Delta. Omicron was the last wave. Therefore, in Fig 2A, the first peak, to the far left (in purple) is the Other variant, next is Beta (in brown), followed by Delta (in green) and the rest (in orange) is Omicron. The number of cases peaked during Beta and Omicron waves.

**Figure 2:**
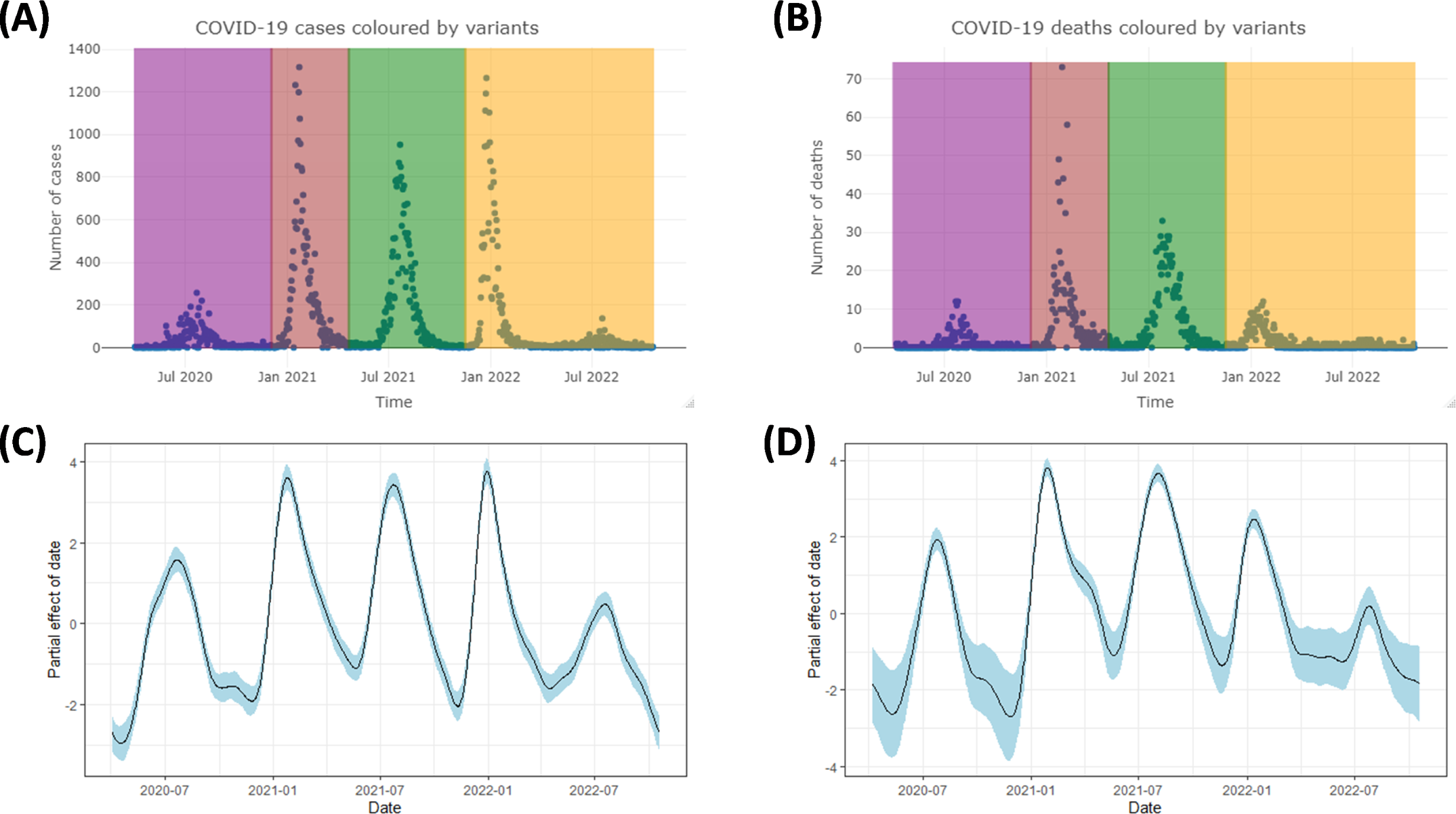
Trends for SARS-CoV-2 cases and deaths. (A) Daily cases coloured by variants, Other(purple), Beta(brown), Delta(green) and Omicron(orange). (B) SARS-CoV-2 Daily Reported Deaths, coloured by variants, Other (purple), Beta (brown), Delta (green) and Omicron (orange). (C) Partial effect of day on cases. (D) Partial effect of day on deaths

Case data is over-dispersed: the variance to mean ratio is 436, which is considered large. The mean is much greater than the median owing to the presence of few large values of reported cases. Results of a GAM model fit to case data using a negative binomial distribution family with a log link function, where cases were a function of day and day-of-week are visualised in Fig 2C. The model plot shows the partial effect of day on the cases and has has five waves; the first three waves are for Other, Beta and Delta, in that order while the last two waves are for Omicron; suggesting that Beta and Omicron were more infectious than Other and Delta variants. The spline in Fig 2C also shows a strong positive relationship between time (day) and cases in the initial phase of the pandemic. There was a decrease in disease transmission during September and October 2020, followed by a mild increase in November 2020 then a decrease again till early December 2020. This marked the end of the first wave, which was dominated by Other variant. The same December 2020, the Beta variant erupted and reached a peak around January 2021 then decreased drastically over the next four months. It did not take long before the Delta variant appeared, which ran through April to November 2021 and had a peak around July. During the Delta wave, cases increased with time between June and July, then decreased afterward. Omicron stayed longer than other variants (11 months), cases fluctuated greatly with time and peaked in January and July 2022.

The model summary results found in Supplementary Material S1 indicate that the smooth term for the time (day) is statistically significant (*p <* 2 *×* 10*^−^*^16^), signifying a clear effect of time whereas the day-of-week predictor is not significant (*p* = 0.882). The model summary confirms that the model has an adequate number of basis functions (*k* = 45), supported by a notable difference between the degree of the curve (4) and the predictor’s effective degree of freedom (35), indicating a good fit. The adjusted R-squared value is 82.5%, which considered high. Model diagnostic plots are shown in Supplementary Material S1.

#### COVID-19 reported deaths modelling

As of 19 October 2022, 2, 683 COVID-19 deaths were reported in Malawi. The first death attributed to SARS-CoV-2 in Malawi occurred five days after the first case was reported. Fig 2B provides the number of daily deaths recorded from 7 April 2020 to 19 October 2022. Maximum and minimum number of deaths observed were 73 and 0, respectively. On average, 3 people died every day due to COVID-19 and the variance was 44.0922. Ninety-five percent of the values for daily new deaths were below 15. Colours in Fig 2B represent different variants. Variant of Other (purple), Beta (brown), Delta (green) and Omicron (orange). Beta caused more deaths than the rest of the variants did. Although Omicron stayed the longest, it caused few deaths, below 14 per day. More than half of the values, (53.5%) are zeros.

Fig 2D illustrates a GAM model fit to death data using a negative binomial distribution with log link function. There were more COVID-19 deaths during the second (Beta) and the third (Delta) waves. Delta was less infectious but caused more deaths, unlike Omicron, which was aggressive but caused few deaths. Deaths increased with time in the initial phase of the pandemic, just like cases did. The decreasing trend took about a month and a half before a strong positive relationship between time and the number of deaths was observed from May to July 2020. Later the relationship became negative till November 2020 aside that it became constant for a moment around October 2020. Deaths during the Beta variant increased rapidly with time from December 2020 and reached a peak around January 2021, then started to drop till May 2021. Deaths during Delta rose to a high point from June 2021 and peaked August 2021, then the trend declined over the next three months before the Omicron deaths were observed starting from December 2021 to October 2022. Omicron deaths had two peaks, in January and August 2022.

The smooth term, day is significant (*p <* 3 *×* 10*^−^*^8^) and the predictor’s effective degrees of freedom are 30. Adjusted R-squared = 76%, which is accepted. The intercept is significant unlike the day-of-week. Diagnostic plots are shown in S1.

### Growth rate and Doubling time

#### Growth rate and doubling time of SARS-CoV-2 cases

Fig 3A shows a growth rate curve for cases with 95% confidence interval. When the growth rate curve for cases is coloured by COVID-19 variants, Fig 3C is produced. The coloured figure allows for comparison of the infection’s growth during different waves, which suggests a rapid increase of the infection during Beta and Omicron as compared to Other and Delta waves.

**Figure 3:**
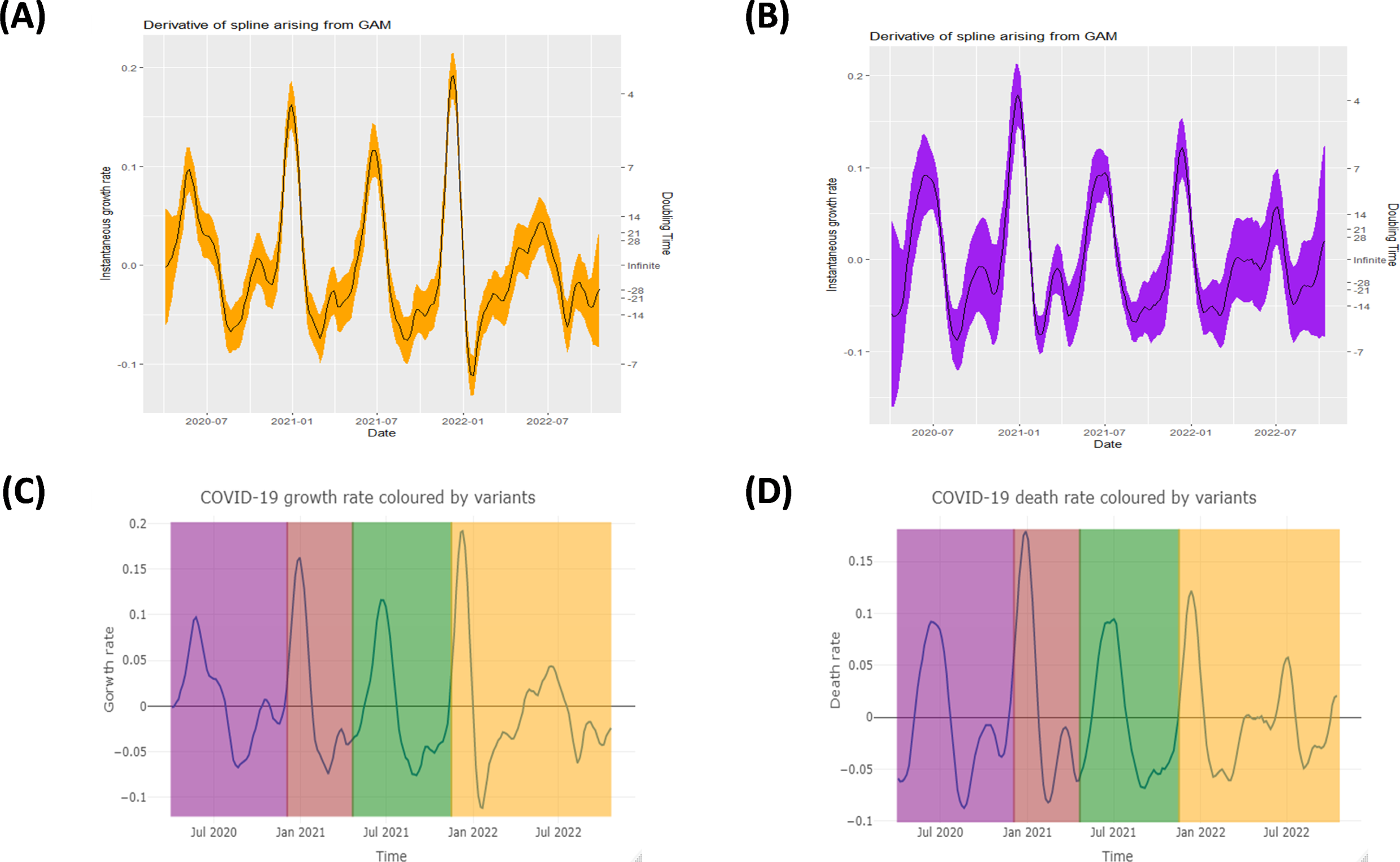
Growth rate and doubling time. (A) Growth rate and doubling time curve for cases. (B) Growth rate and doubling time curve for deaths. (C) Growth rate for cases: Other (purple), Beta (brown), Delta (green) and Omicron (orange). (D) Growth rate for deaths: Other (purple), Beta (brown), Delta (green) and Omicron (orange).

#### Growth rate and doubling time of SARS-CoV-2 deaths

As shown in Fig 3B, the growth rate for deaths from 7 April 2020 to 19 October 2022 varied between *−*0.09 and 0.185 per day, not very different from growth rate for confirmed cases. Unlike in reported cases, lowest and highest growth rates for deaths were observed during Other and Beta variants, respectively. Negative growth rate and doubling time indicate that cumulative deaths were halving; and doubling otherwise. The general growth rate trend for deaths appears to be similar to that of cases. Fig 3D shows death rate curve coloured by variants.

### COVID-19 Reported Cases by District

SARS-CoV-2 confirmed cases across the regions in Malawi are visualised in Fig 4, where Fig 4A shows total confirmed cases, Fig 4B illustrates total population and Fig 4C the case incidence per 1,000 people. Blantyre city in the southern region had the highest number of reported cases, 24, 189 followed by the capital city, Lilongwe which had 19, 822 cases. The least number of total confirmed cases, 118 was observed in the northern region, particularly Likoma island. Three-quarters of the districts realised total cases below 2, 016. About 64.6% of the total confirmed cases were observed in the four major cities of Malawi including Blantyre, Zomba, Lilongwe and Mzuzu.

**Figure 4:**
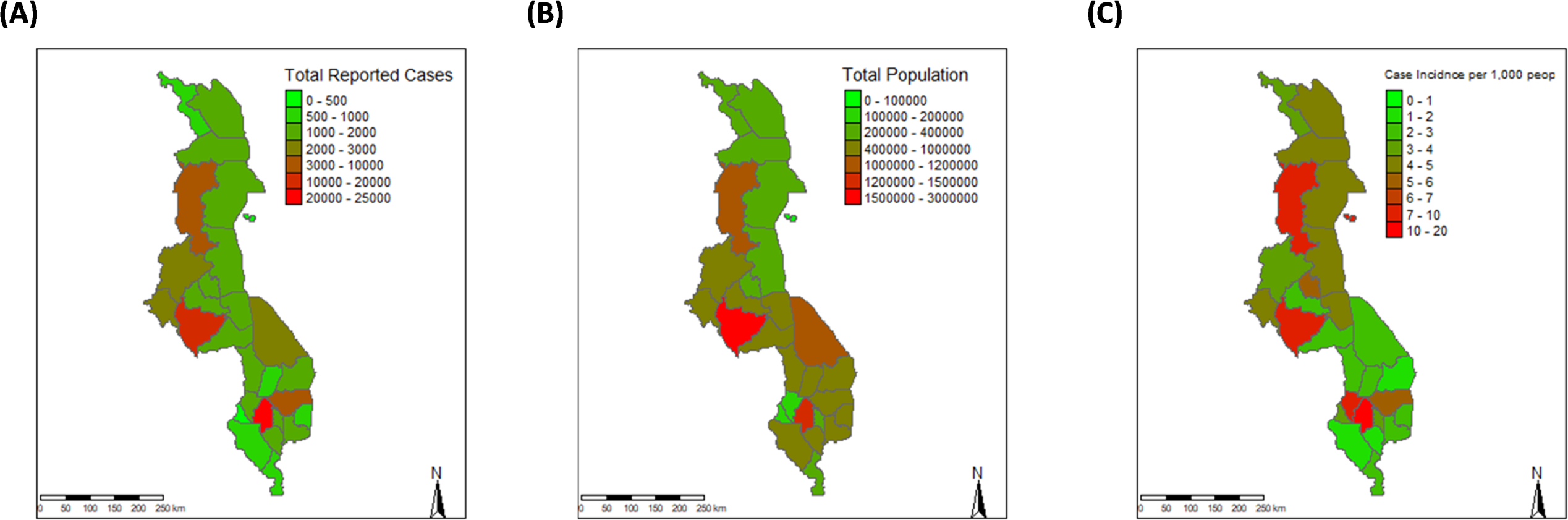
Maps of confirmed cases, total population and cases per capita. (A) Total confirmed cases in districts. (B) Total population in districts. (C) Case incidence per 1, 000 people: Likoma Island and the major cities; Mzuzu (Mzimba), Lilongwe, Blantyre and Zomba have high incidence.

### COVID-19 phylogenetics

#### Spread of COVID-19 variants across the regions

More than half of the sequences, 57.3% used in this study were collected from the southern region while 10.2% from the central and 5.8% from the northern part of Malawi however, there was missing information on the region for 26.6% of the sequences. When grouped according to COVID-19 variants as described by WHO, the majority of the sequences belonged to Delta (39.63%), followed by Beta (34.93%), and then Omicron (19.67%). Other and Alpha variants had the least number of sequences, 5.35% and 0.42%, respectively. A time tree that resulted from the phylogenetic analysis is shown in Fig 5, where clusters of sequences representing different variants are noticeable. All variants evolved in 2020 however, Omicron took longer to mutate into sub-variants after after first emergence than the rest. Comparing the five variants, Beta is closer to Delta and both are closer to Alpha than they are to Omicron. Alpha did not circulate much as depicted from the phylogeny. Fig 5 also shows that Omicron mutated into sub-variants BA.1 and BA.2 where from BA.2 three other sub-variants, BA.4, BA.5 and BQ evolved.

**Figure 5:**
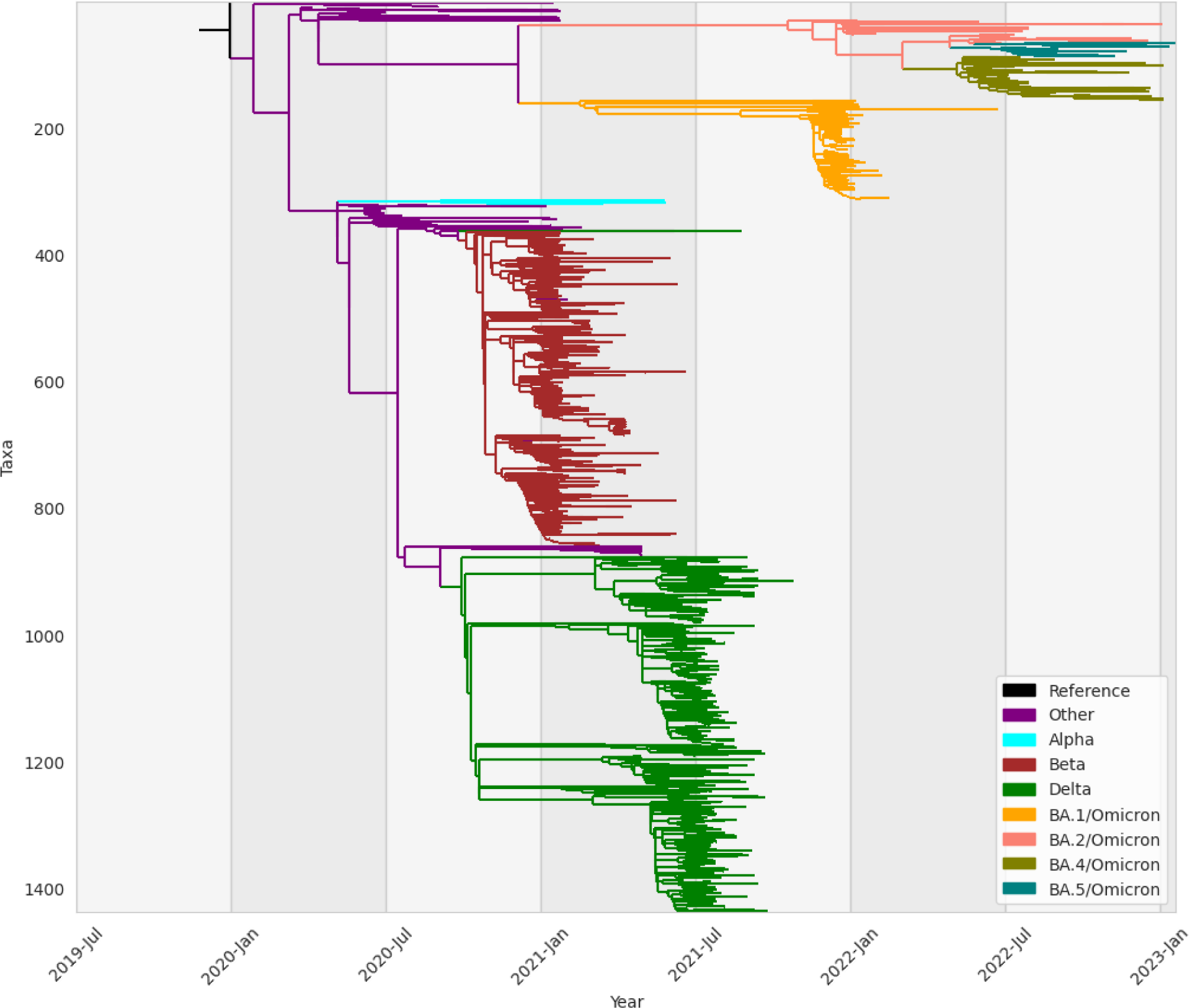
Maximum likelihood phylogenetic timetree. Branches coloured according to SARS-CoV-2 Variants: Alpha (blue), Other (purple), Omicron (orange, salmon, olive, teal and magenta), Delta (green) and Beta (brown)

The distribution of SARS-CoV-2 variants across the regions of Malawi is shown in the table in S1. Beta, Delta, Omicron and Other variants spread across all three regions of Malawi while it is hard to conclude for Alpha because of missing information. However, Alpha, Delta and Omicron were more dominant in the southern region.

Phylogenetic analysis of each COVID-19 variant revealed that Alpha in Fig 6A, Delta in Fig 6C and Omicron in Fig 6D were more common in the southern region, which is attributed to high population density in the region. Population density is high in the southern region possibly due to the presence of a major commercial city, Blantyre. However, it was not obvious for Beta in Fig 6B and the variant of Other in Fig 6E to identify the regions they were common because 40% and 60% of their sequences respectively, had missing information on the region they were collected from. Despite the absence of information for the majority of the sequences in some variants, it can be observed from the individual trees that there are possibilities for sequences to be close but found in different regions. Not all related sequences were found in one region i.e. a genome of a sequence found in the North could be very close to another one in the South.

**Figure 6:**
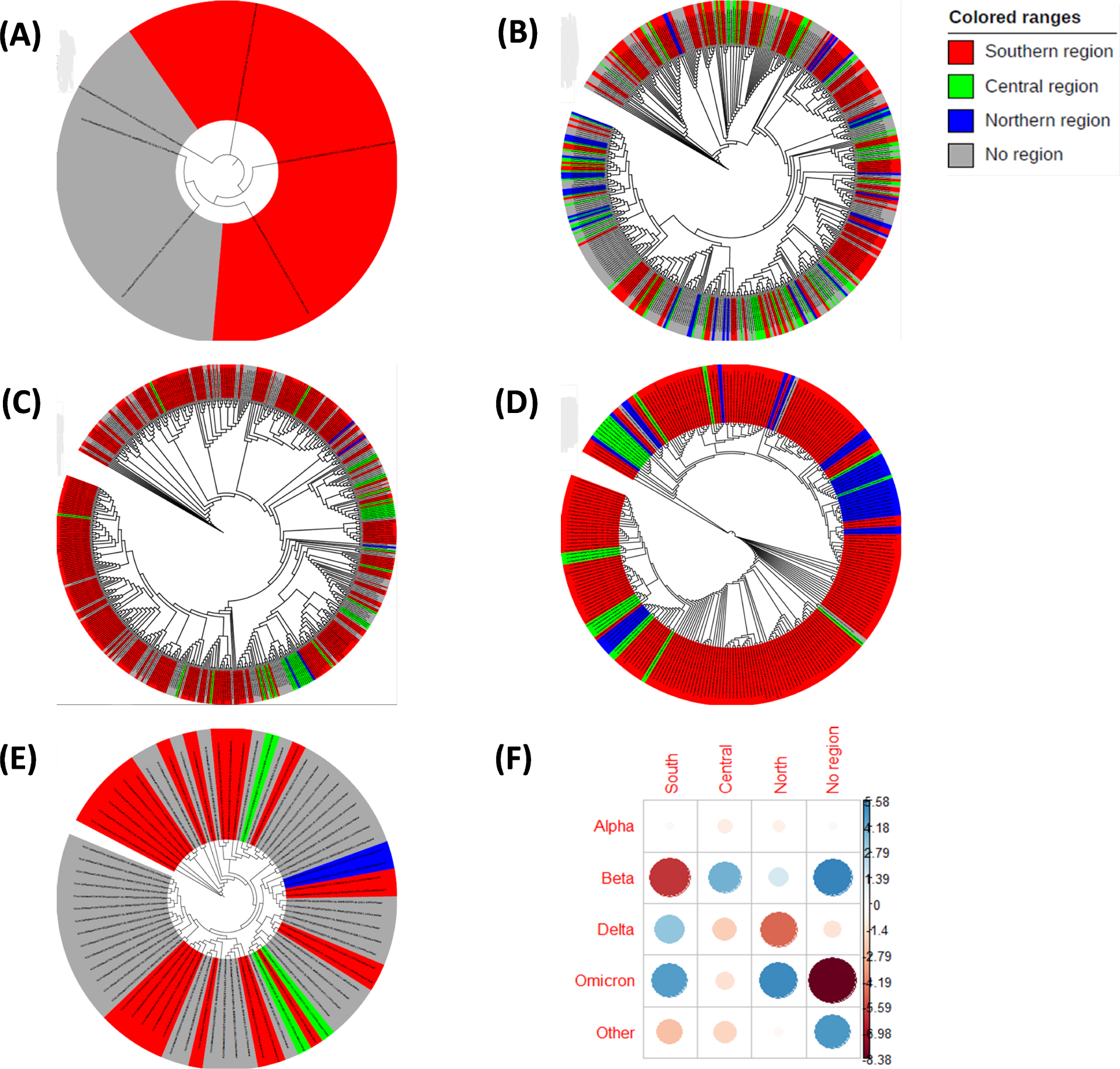
COVID-19 variants and geographical heterogeneity. (A) Alpha variant. (B) Beta Variant. (C) Delta variant. (D) Omicron variant. (E) Other variant. (F) Pearson residuals

#### Chi-square test for the difference in proportions of COVID-19 variants in the regions

Proportions of COVID-19 variants across the regions and their confidence intervals (CI) are reported in a table in Supplementary Material S1. The CI for Alpha could not be computed accurately due to the small number of samples. The contingency table for the spread of COVID-19 variants across the three regions is visualised in the figure found in Supplementary Material S1. The *χ*^2^*−*test for the difference in proportions found that the proportions of SARS-CoV-2 in the regions were significantly different (*p ≈* 0.0005).

Further analysis of the residuals of the test revealed the cells that contributed to the magnitude of the *χ*^2^*−*score. As indicated in Fig 6F, with a cut-off point of *|*3.02*|*, Beta sequences are significantly less in the southern region than expected however, significantly more than expected in the central region. Delta is significantly more (less) than expected in the southern (northern) region, and Omicron is significantly more than expected in the southern and northern regions. Other variant is significantly less in the southern region than expected. Rated out of 100, cell contribution to the *χ*^2^ score is visualised in Supplementary Material S1. It is clear that Beta in the southern region and that with missing information contribute greatly towards the *χ*^2^ score. Sequences of Other without region information and Omicron in the noth and south also contribute highly to the score. This implies that the pandemic hit the northern and southern regions more than the central during the Omicron wave while the southern and northern regions were the least affected during the Beta and Delta waves respectively.

## Discussion

The far-reaching consequences of COVID-19 throughout the world as well as individual states motivates the need for extensive research around the pandemic. Many countries were affected in various sectors such as health, social, economic and many others. The panic was intense in low and middle-income countries considering that their resources already are limited[23]. It was, therefore, worth understanding the disease dynamics, the evolution of the virus and the burden the pandemic posed on a low-income country like Malawi to aid in the formation of policies and interventions at present or in the future. The study analysed epidemiological and phylogenetic publicly available data of SARS-CoV-2 in Malawi from the day the first case was identified to 19 October 2022 and 19 January 2023, respectively. With the use of generalised additive models, te infection’s trend and groethrate were described while the use of IQtree, TreeTime and iTOL allowed the capturing of the times of the most recent common ancestors for the variants as well as the geographical distribution of the virus.

The study finds a case fatality rate of 3% (2.8% to 3.1% credible interval) as of 19 October 2022, which is higher than a 2.4% CFR as of 1 September 2020 that Dalal (2021) found in their study that involved twenty African countries in the WHO region excluding Malawi [42]. Results show that Malawi encountered five waves of SARS-CoV-2 (see Fig 2A), and that each wave was dominated by one variant as the time tree, as Fig 5 illustrates. Other variants evolved first, followed by Beta then Delta. Omicron emerged last. This chronological order agrees with the literature particularly study results of Anscombe (2023)[43]. In their study conducted between July 2020 to March 2022 in Malawi, in which they compared the waves, Anscombe (2023) [43] did not find Alpha variant, which is not surprising as our study has found that the proportion of alpha was as low as 0.4%, and spread during the times Beta and Delta dominated.

Comparing the waves in terms of cases, the study found that the first wave, Other was the least infectious of all as seen in Fig 2C and Fig 3C, but this could be due to low testing or under-surveying as it was the early stage of the pandemic. Vaccination was launched at the time Beta was phasing out and Delta emerging[44] and results show that Delta was less infectious compared to Beta. This minimal infectiousness of Delta could be attributed to the effect of the vaccine as vaccination triggers herd immunity hence, the transmission of an infection from infected to susceptible individuals is blocked[45]. However, Omicron was more aggressive despite that a good proportion of the population had been vaccinated and that herd immunity was high. Although Omicron was more infectious and highly transmissible it caused fewer deaths than Beta and Delta (see Fig 2D and Fig 3D), which agrees with results of the study by Butt (2022) [46] in Qatar and Katella (2022) [10]. Similar results are obtained in this study as shown in the figure demonstrating the posterior distribution of CFR found in S1, which suggests that Omicron has the lowest CFR while Delta has the highest CFR followed by Beta.

The *χ*^2^ test showed that proportions of COVID-19 variants across the regions of Malawi were significantly different implying that the variants posed unequal impact on the regions. Precisely, Omicron was worse in the southern and northern regions compared to the central region. Beta did not affect the southern region much, as Delta did in the north.

Disease incidence was relatively high in the four cities of Malawi (see Fig 4), most likely because they are commercial areas where the majority of businesses take place. Two of the cities lie in the southern region, one is found in the central and the other one is in the north. Increased disease incidence in Likoma Island is owed to low population. Phylogenetic trees for individual variants showed that close sequences were found in different regions, indicating there was inter-regional mobility of people although travel was restricted.

## Conclusion

Various lineages of SARS-CoV-2 were found in Malawi these include A.23.1, B, B.1, B.1.1.1, B.1.1.412, B.1.1.7, B.1.351, B.1.36, AY.1, AY.3, AY.4, B.1.617.2, B.1.617.3, BA.1, BA.2, BA.4, BA.5 and BQ.1. Five variants according to WHO naming were formed from these lineages; Alpha, Beta, Delta, Omicron and the first lineages that evolved before the variants of concern (VOC), which in this study has been constantly referred to as ‘Other’ variant. Of the five variants, Delta and Beta had higher percentages, 39% and 34% respectively. Alpha had the lowest proportion of 0.4%. The five waves of the pandemic Malawi encountered were of Other (non-VOC), Beta, Delta and the last two were Omicron, in that order. The disease burden, which was measured through the case fatality rate was found to be relatively lower compared to high-income countries but relatively higher when compared to other low-income countries. Cases and deaths increased around January, June, July or December however, the infection had a general decreasing trend. This implies that in the future, if SARS-CoV-2 or similar flu infections emerge, more attention will have to be paid during these months when transmission is high. Additionally, more care should be taken with Omicron as it is more infectious and Beta because it kills faster. Factors associated with low disease severity observed in the early phase could be but not limited to low testing, early implementation of preventive measures; and others beyond these such as climatic differences, a high proportion of the younger population, pre-existing immunity and genetics. COVID-19 vaccination also played a role in combating the pandemic as this was noticed in the reduced growth rate of the infection soon after vaccination was introduced, the time Delta variant emerged. Proportions of the variants across the three regions of Malawi were significantly different, indicating a variation in the burden posed by the pandemic in the regions during different waves. Omicron affected the north and southern regions most while Beta and Delta affected the southern and northern regions respectively, less than expected. Therefore, northern and southern regions need to be prioritised as far as resource allocation to combat SARS-CoV-2 and other related infections is concerned. DNA mutations that occurred in the variant of Other resulted in the remaining four variants; Alpha, Beta, Delta and Omicron. Three variants, Alpha, Beta and Delta are close as they share recent common ancestors. These results agree with what other researchers found, especially on the chronological order in which the variants evolved. It is also worth noting that the genetics of SARS-CoV-2 change frequently within a short period causing rapid divergence of sequences, which leads to numerous lineages or variants.

## Code availability

Code for the manuscript is available at https://github.com/MwandieKambaAfuleni/Covid19-Malawi.

## Supporting information

**S1 Supplementary Material**

## Data Availability

The manuscript presents secondary analysis of public data. We are not the data
guardians for this, so provide sources in the manuscript to allow other researcher to
obtain the data from its owners.

https://gisaid.org/

https://ourworldindata.org/

## Acknowledgments

MKA was supported by the Schlumberger Foundation–Faculty for the Future. KL was supported by the Royal Society and the Wellcome Trust (107652/Z/15/Z) and the Li Ka Shing Foundation. OJ was supported by the Wellcome Trust (228264/Z/23/Z). KL, IH and TH were supported by the Wellcome Trust (227438/Z/23/Z). RC, IH and TH were supported by the UKRI Impact Acceleration Account (IAA 386).

We gratefully acknowledge all data contributors, i.e., the Authors and their Originating laboratories responsible for obtaining the specimens, and their Submitting laboratories for generating the genetic sequence and metadata and sharing via the GISAID Initiative, on which this research is based.

---

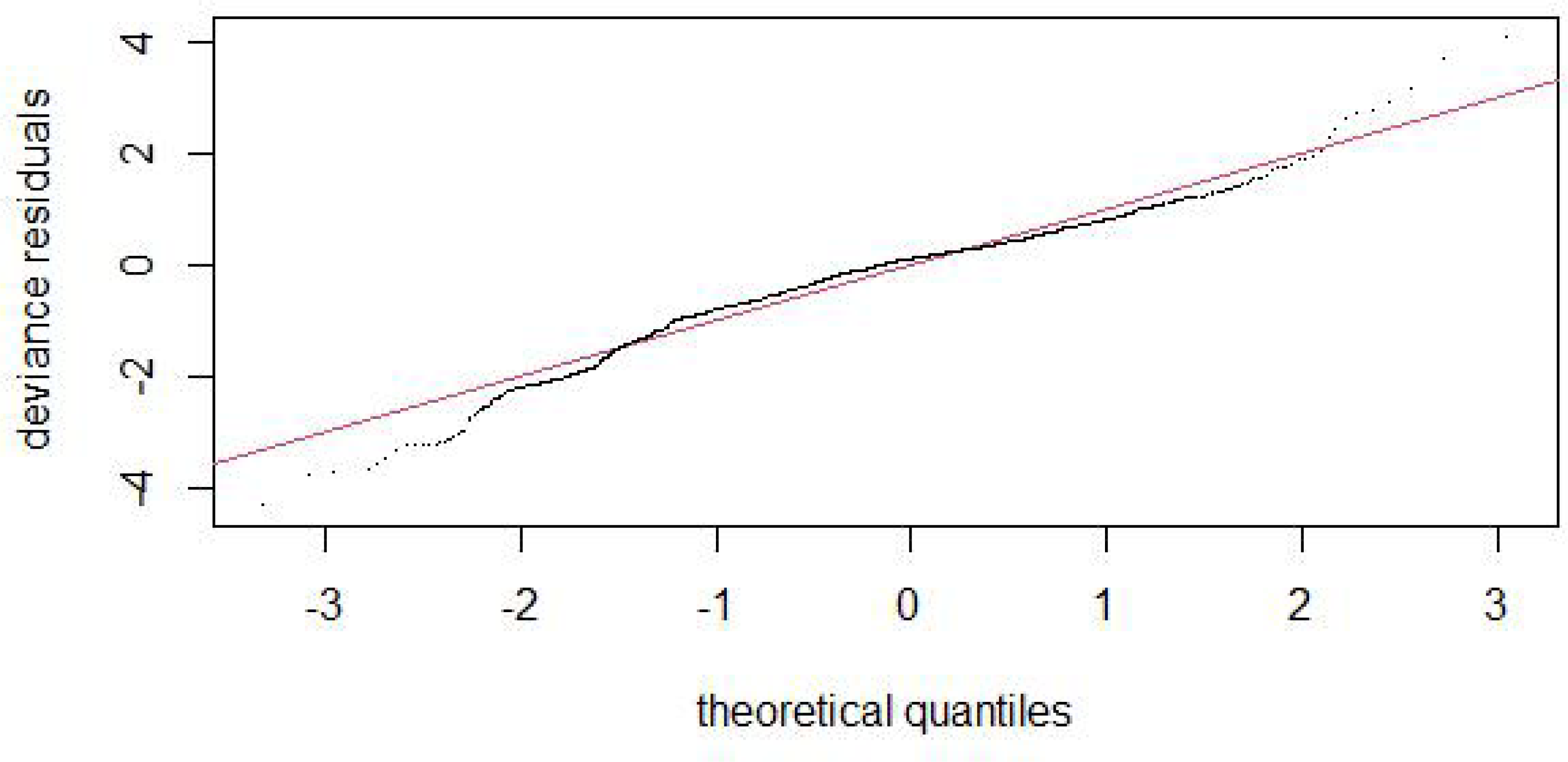

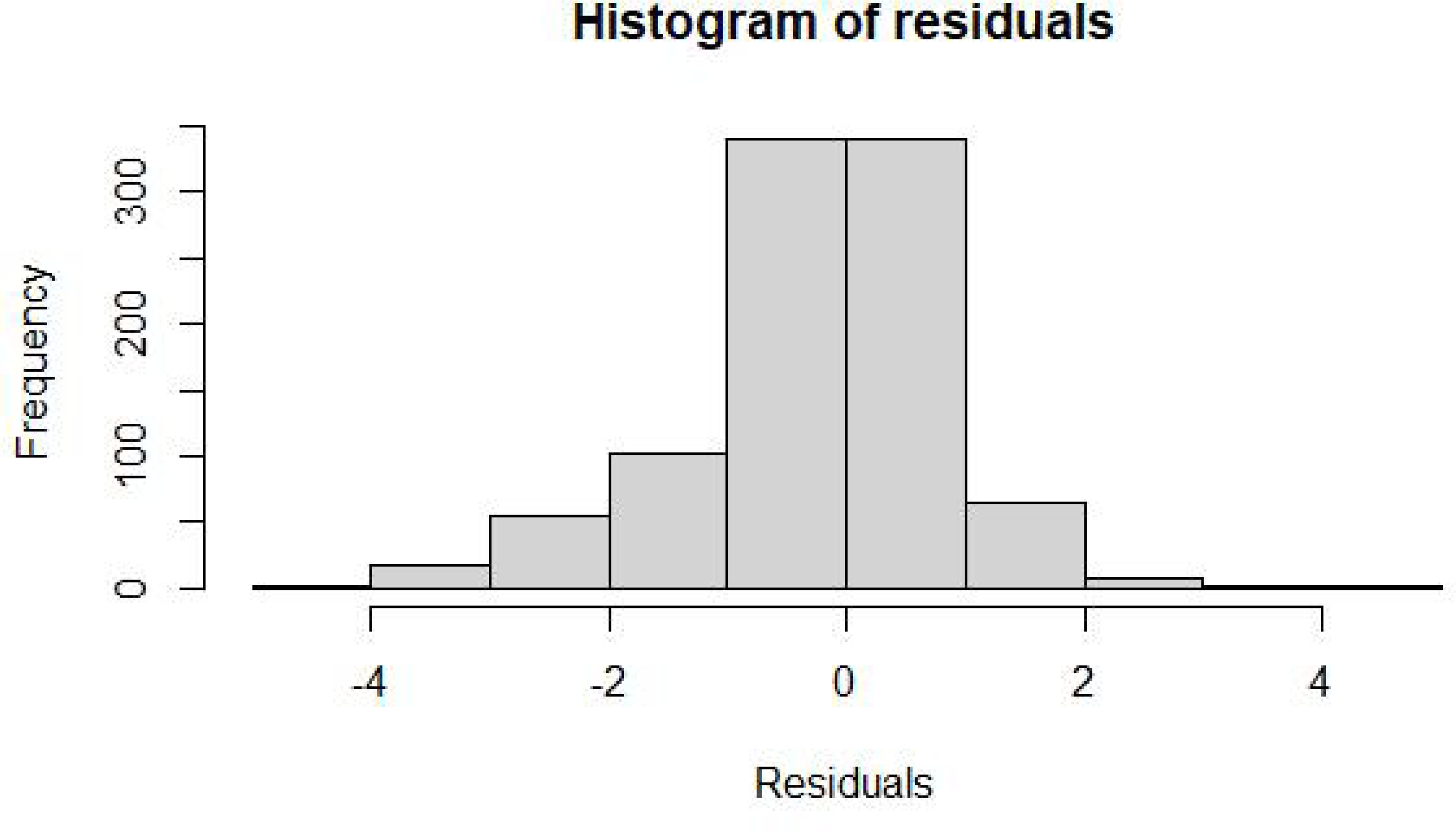

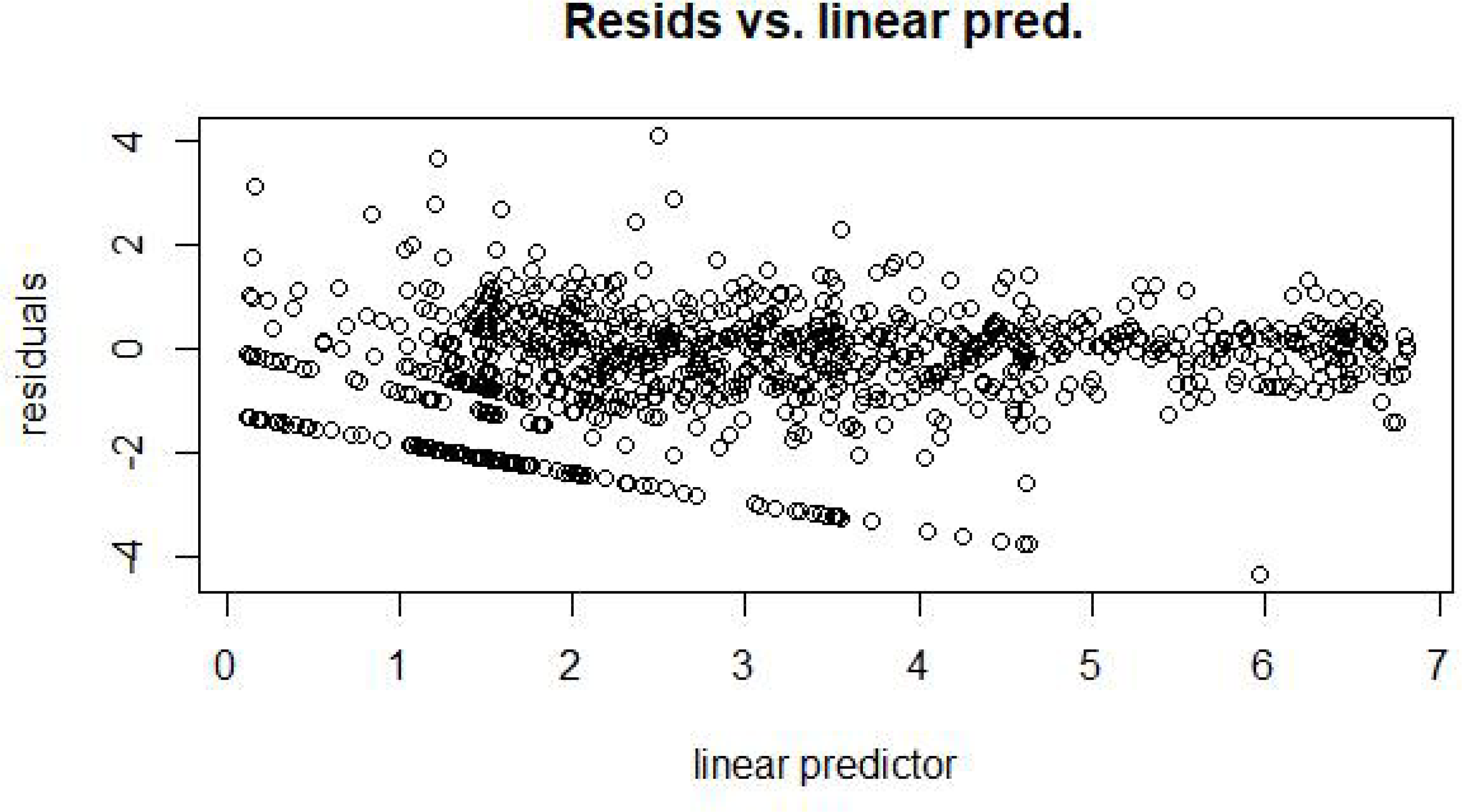

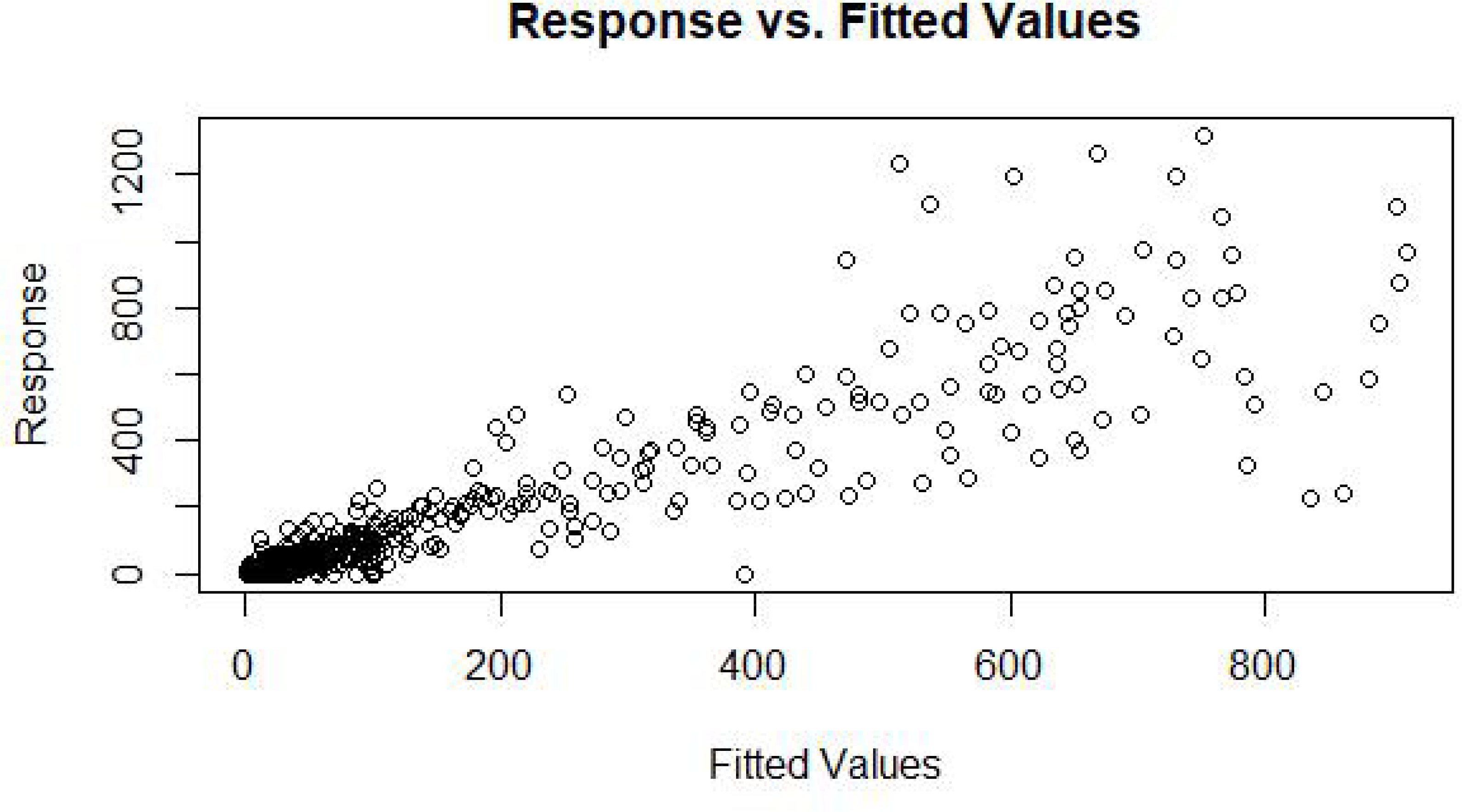

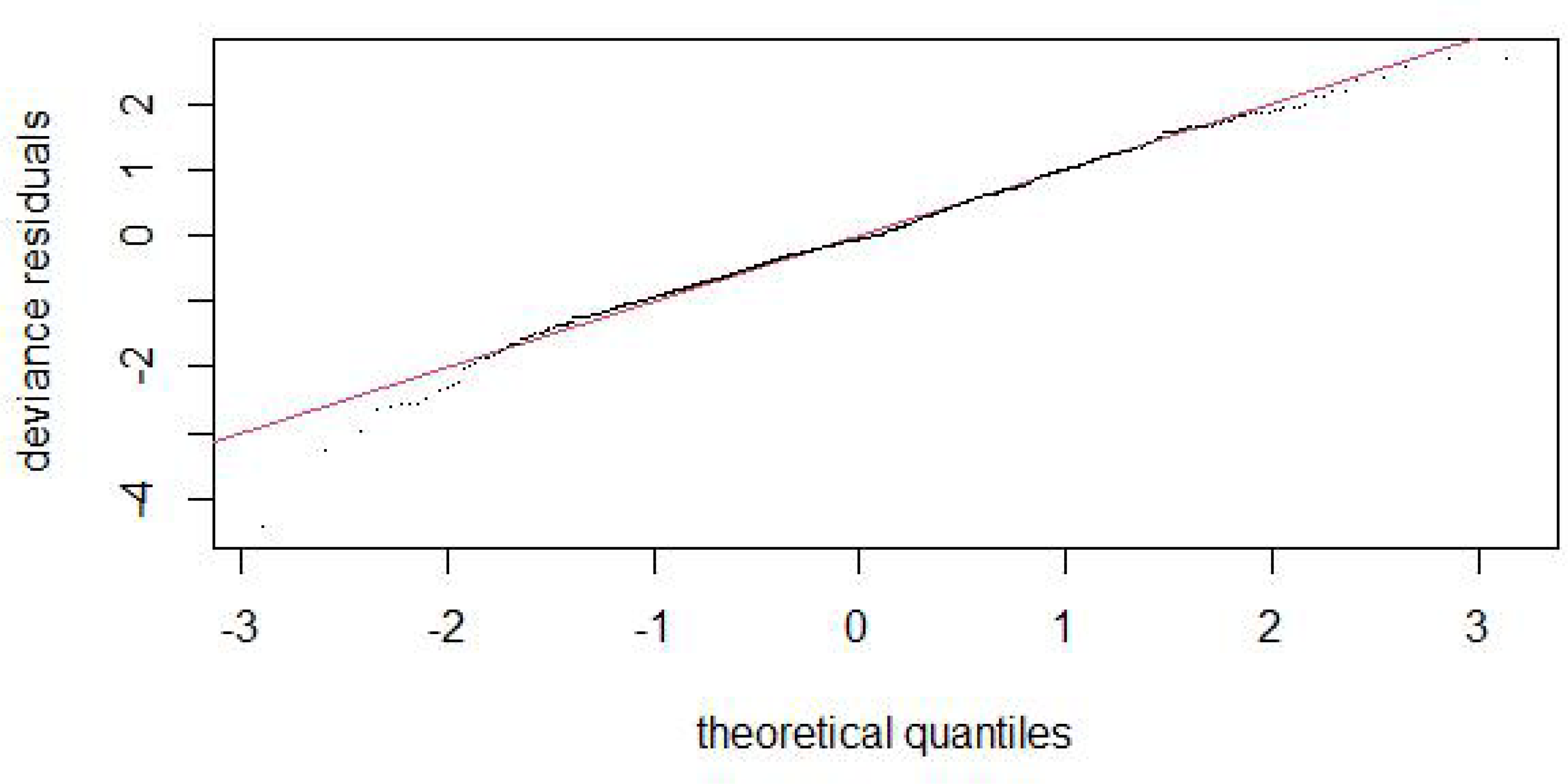

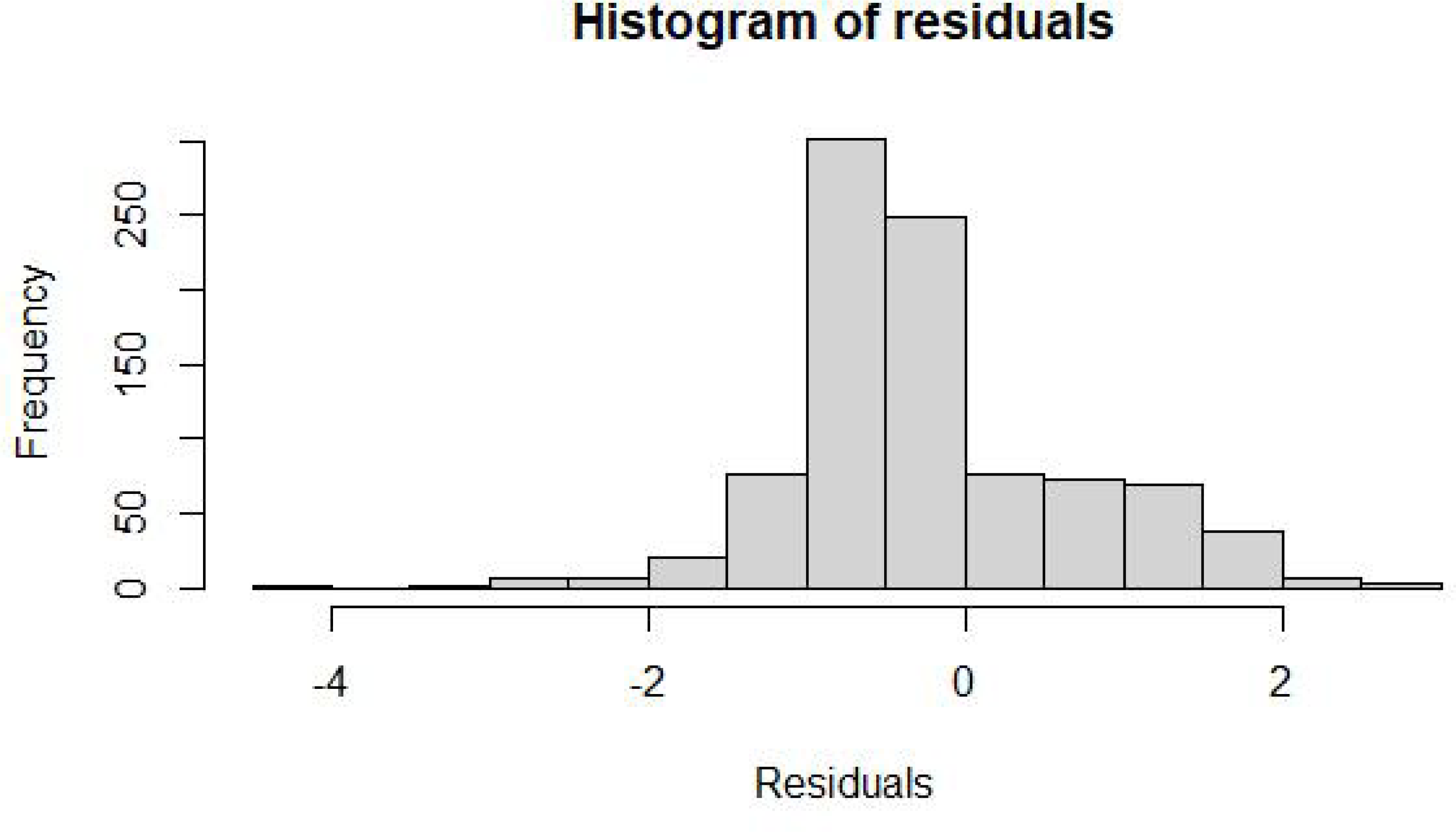

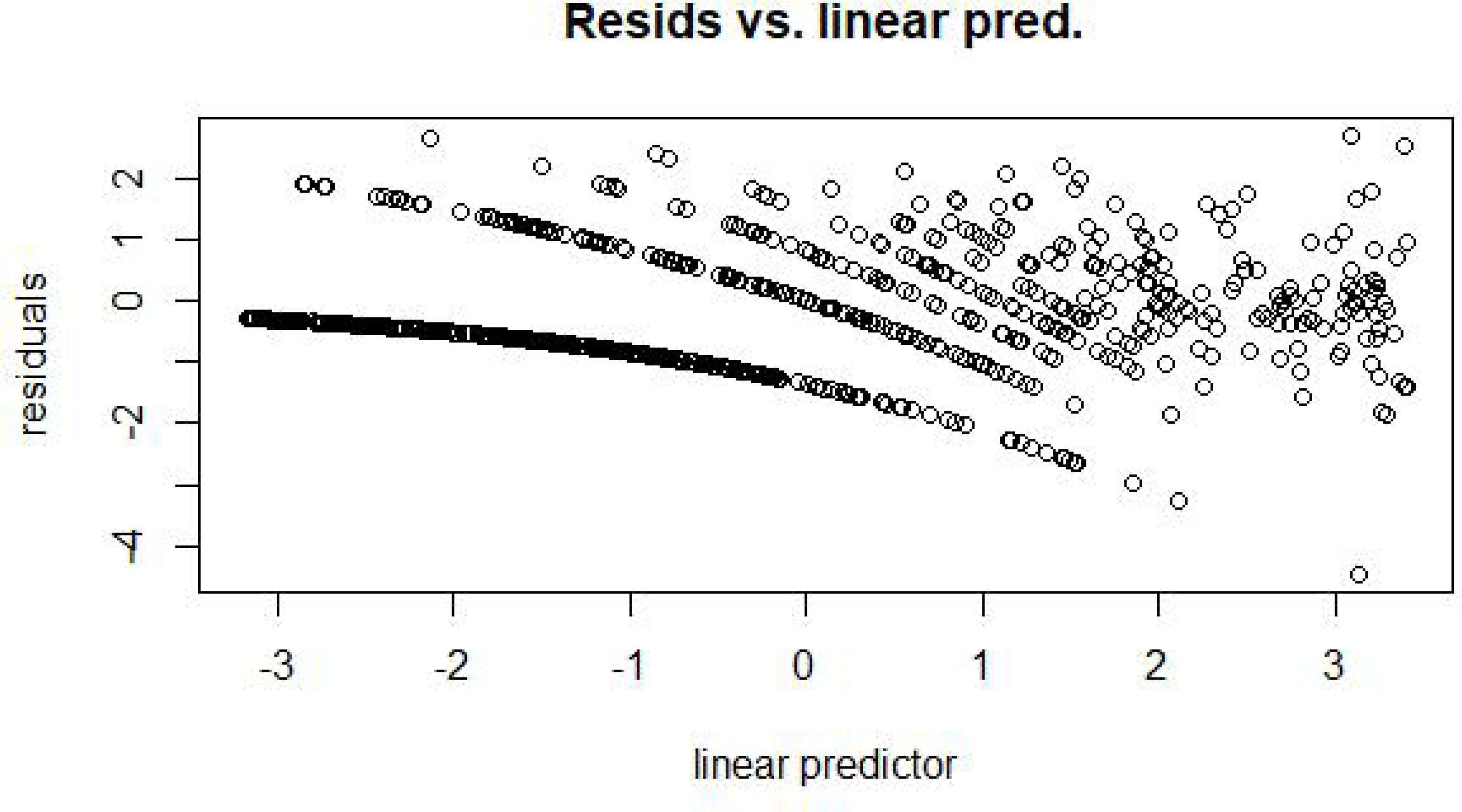

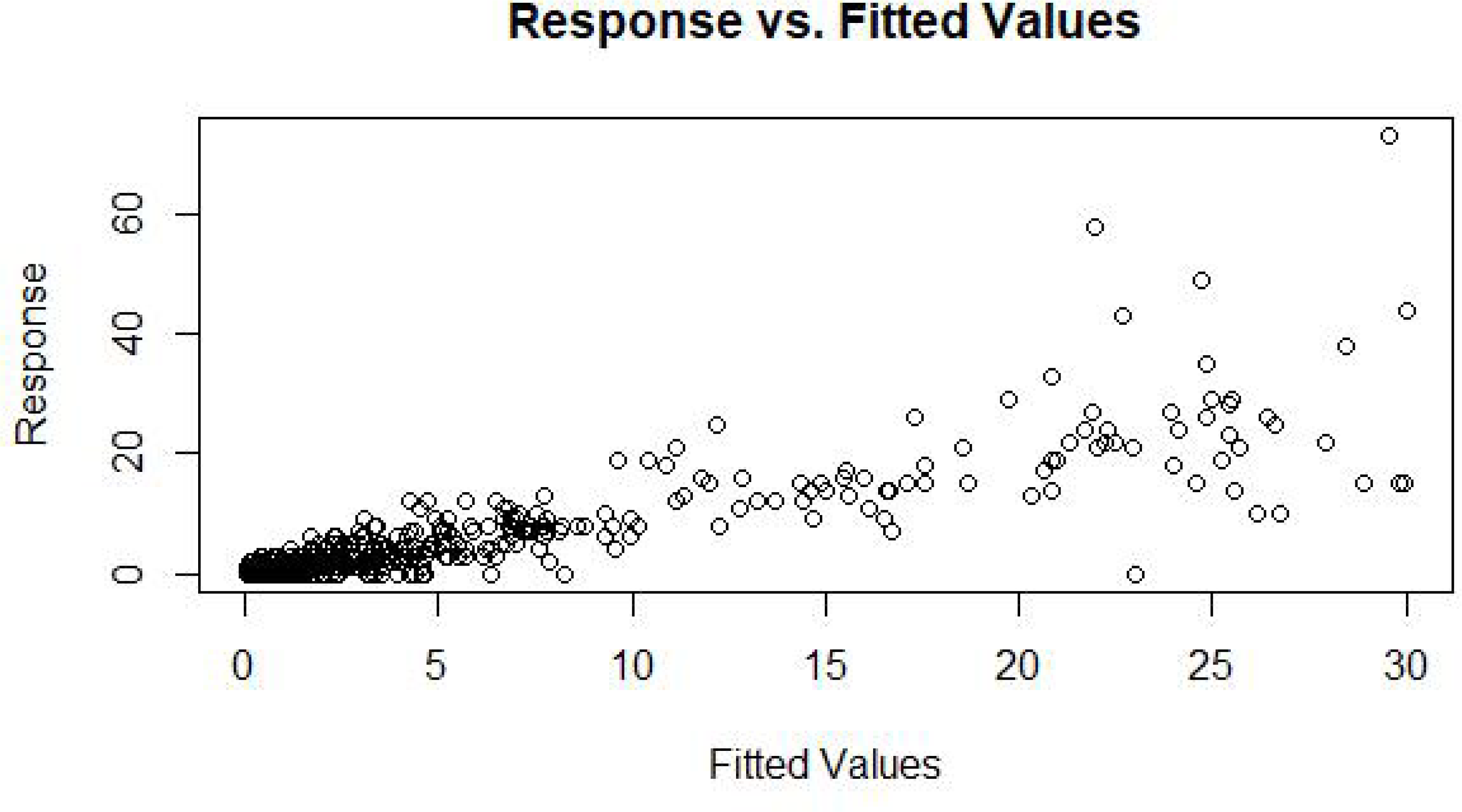

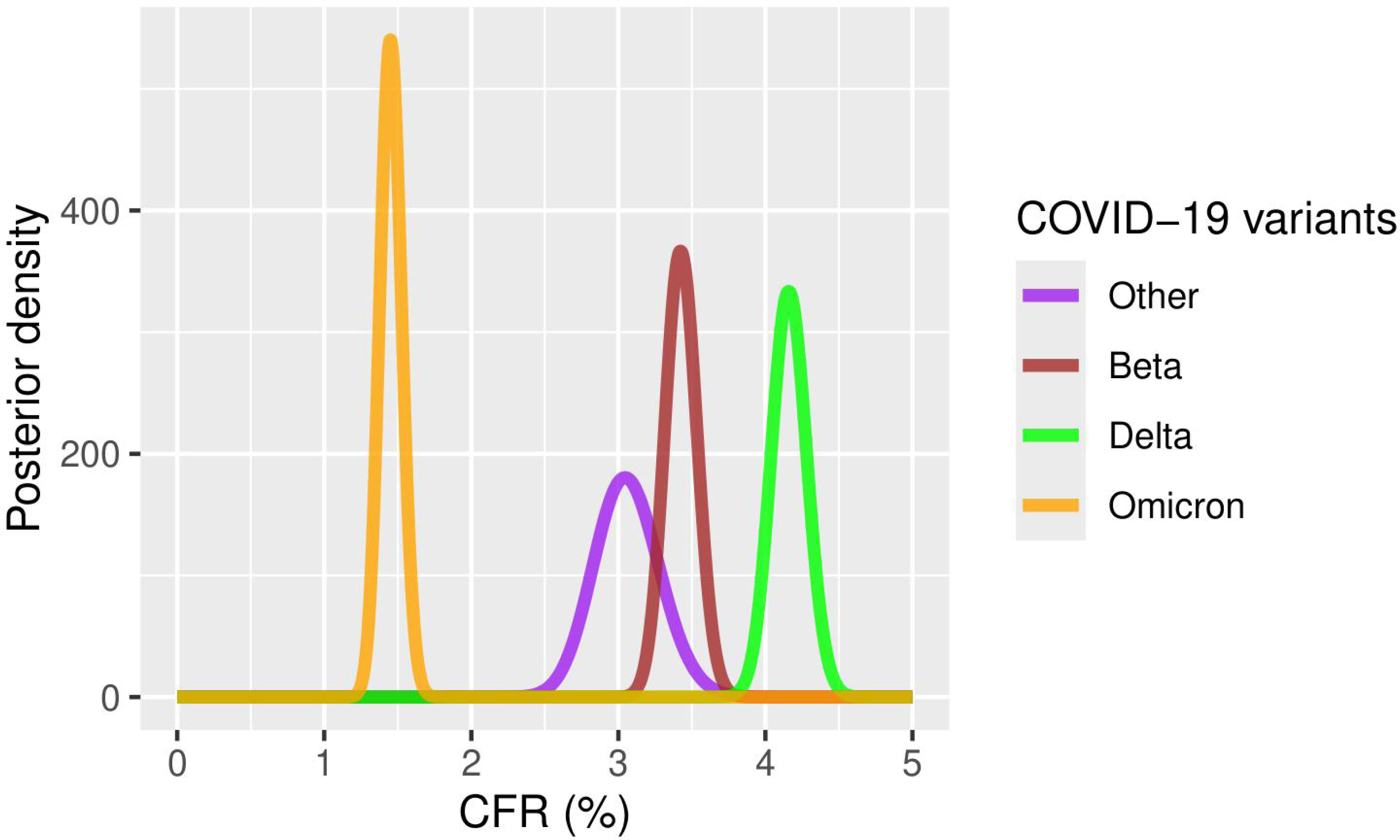

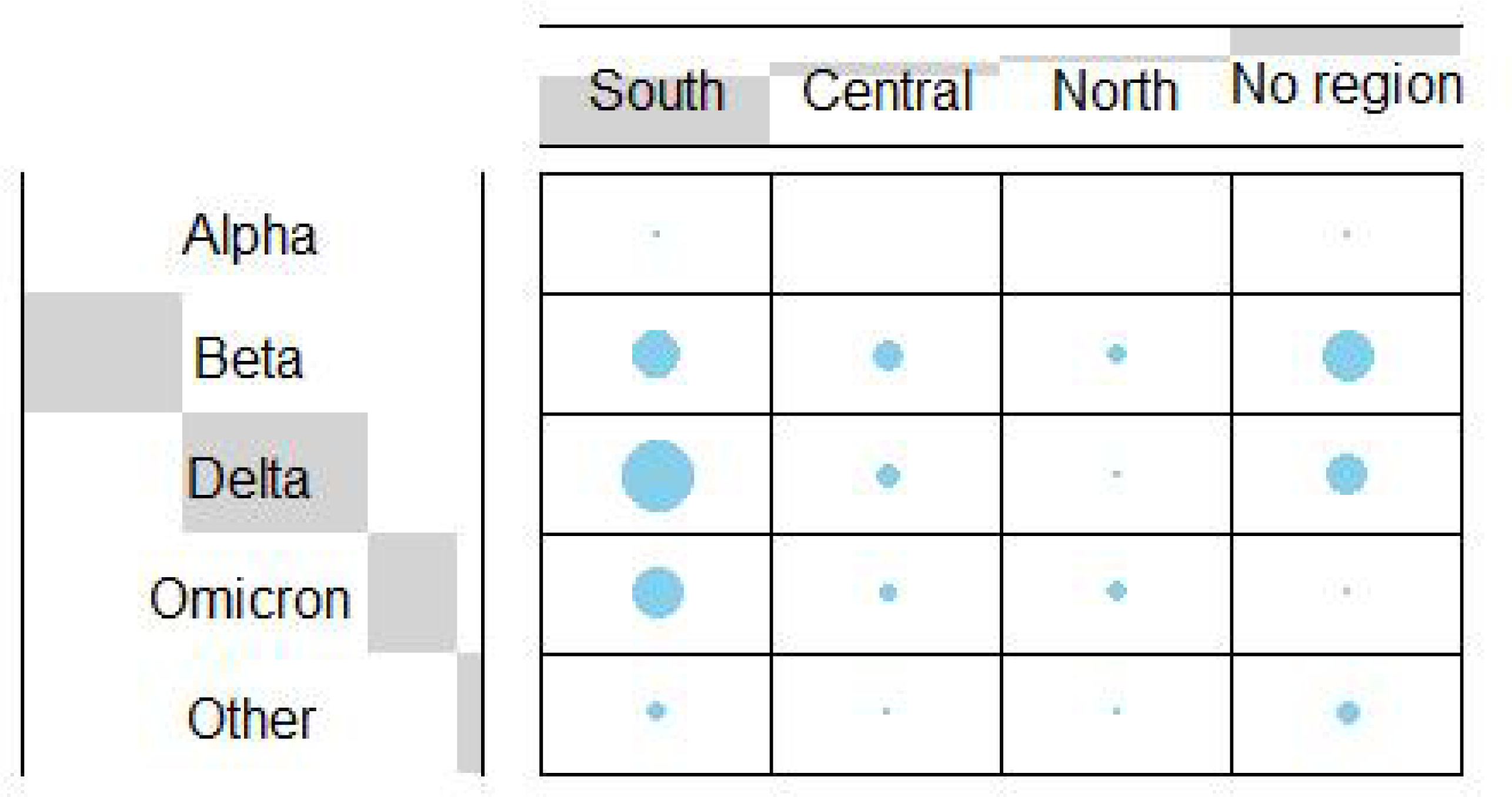

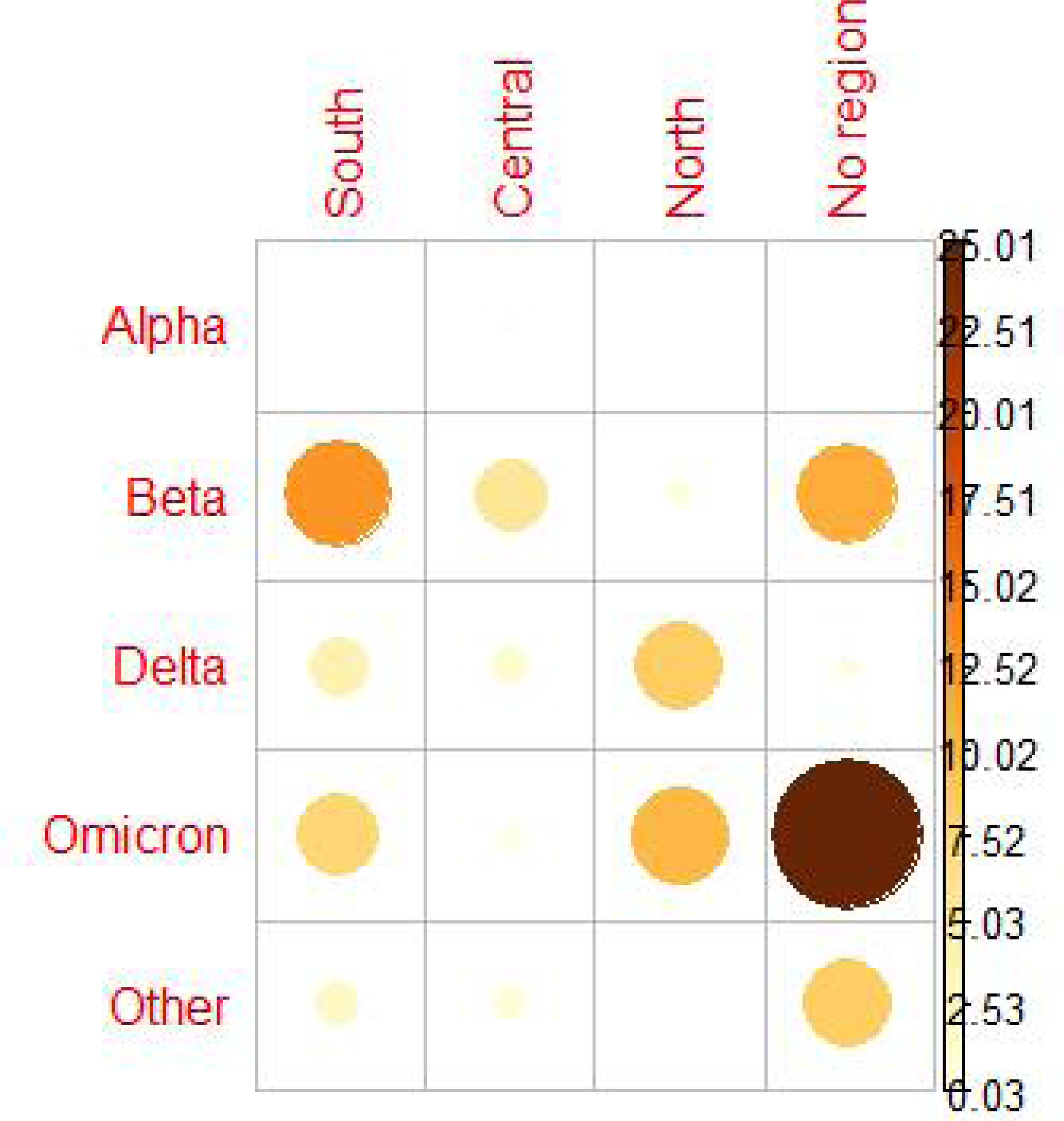

