## Supplementary Material for "Epidemiological and phylogenetic analyses of public SARS-CoV-2 data from Malawi"

### Supplementary material for the Epidemiological and phylogenetic analyses of public SARS-CoV-2 data from Malawi

#### 1 Data analysis

##### 1.1 Generalised Additive Models (GAM)

Implemented in R using the mgcv package, generalised additive models with the negative binomial family were fitted to data to describe trends of cases and deaths over time. Two fixed covariates, day and day of week were considered and the corresponding model smooth term of class s was defined for the day explanatory variable; with a cyclic cubic regression spline (bs = "cc") and enough basis dimension ($k = 45$). Below is the model,

$$g(\mu_i) = \mathbf{A_i}\theta + f_1(x_{1i}) + f_2(x_{2i})$$

where $\mu_i \equiv \mathbb{E}(Y_i)$ and $Y_i \sim \mathbf{EF}(\mu_\mathbf{i}, \phi)$ is a response variable (cases or deaths) with an exponential family distribution (negative binomial) with mean $\mu_i$ and scale parameter $\phi$. $\mathbf{A_i}$ is the row of a model matrix, $\theta$ is the parameter vector, $f_1$ and $f_2$ are smooth functions of covariates, $x_1$ and $x_2$. A smooth function is denoted by;

$$f(x) = \sum_{j=1}^{k} b_j(x)\beta_j$$

where $\beta_j$ represents the value of unknown parameter and $b_j$ is the basis of expansion (i.e $x^0$, $x^1$, $x^2$, $x^3$ e.t.c) [1].

Negative binomial and quasi-poisson distributions are widely used in epidemiology for overdispersed count variables where variance is greater than the mean [1]. Negative binomial assumes that variance is a quadratic function of the mean while quasi-poisson model assumes that variance is a linear function of the mean. In this study, the variance to mean ratio of both case and death data was high, 435.5 and 15.2 respectively, implying that data was overdispersed therefore, negative binomial or quasipoisson family distribution was ideal. However, negative binomial family distribution was chosen because research shows that it performs way better with real-world data than the others because they give a close fit, especially for data that contains $< 30\%$ of significant zero counts [2].

##### 1.2 Growth rate and Doubling time of SARS-COV-2

Occurrence of new cases in a period, the incidence of a disease is denoted by;

$$y(t) = y_0 \mathrm{e}^{rt} + \text{noise}, \qquad (1)$$

where $r$ is growth rate and $y_0$ is the disease incident at time $0$. Number of cases or deaths at time $t$, $y(t)$ is proportional to $\mathrm{e}^{s(t)}$;

$$y(t) \propto \mathrm{e}^{s(t)}$$

where $s(t)$ are smooth functions. The time derivative of the smooth functions returns the instantaneous growth rate, $r = \dot{s}(t) = \dot{f}_1(x_1) + \dot{f}_2(x_2)$ where $x_1$ and $x_2$ are day and day-of-week, respectively [3] [4].

$$T_D = \frac{\ln(2)}{\dot{s}(t)} \tag{2}$$

##### 1.3 Phylogenetic analysis

**Table 1.** SARS-Cov-2 lineages grouped into variants and named according to names given by the World Health Organisation (WHO)

| COVID-19 variant | | | | |
|---|---|---|---|---|
| Alpha (6 sequences) | Beta (492 sequences) | Delta (569 sequences) | Omicron (273 sequences) | Other (82 sequences) |
| B.1.1.7 | B.1.351 | AY.104 | BA.1 | A.23.1 |
| | B.1.36 | AY.116 | BA.1.1 | B |
| | | AY.122 | BA.1.13 | B.1 |
| | | AY.30 | BA.1.14 | B.1.1 |
| | | AY.37 | BA.1.14.2 | B.1.1.1 |
| | | AY.38 | BA.1.17 | B.1.1.33 |
| | | AY.45 | BA.1.17.2 | B.1.1.375 |
| | | AY.46 | BA.1.18 | B.1.1.412 |
| | | AY.6 | BA.1.19 | B.1.1.448 |
| | | B.1.617.2 | BA.1.21 | B.1.1.54 |
| | | | BA.1.9 | B.1.177 |
| | | | BA.2 | B.6 |
| | | | BA.2.65 | |
| | | | BA.4 | |
| | | | BA.4.1 | |
| | | | BA.4.1.9 | |
| | | | BA.4.6.1 | |
| | | | BA.4.7 | |
| | | | BA.5 | |
| | | | BA.5.2 | |
| | | | BQ.1.1 | |

The collected sequences in fasta format were aligned to identify regions of similarities. The sequence alignment process used MAFFT, a multiple alignment program for amino acid or nucleotide sequences, which we coded in python-jupyter notebook. The aligned sequences together with the reference sequence of Wuhan city, China, were later processed into evolution trees using IQ-TREE, a stochastic algorithm that builds phylogenetic trees by the maximum likelihood method [5]. IQ-TREE, which is an efficient, open-source software for phylogenomic inference was chosen for its efficiency in selecting the best model for tree reconstruction. The tree was rooted with a reference sequence of Wuhan using a model $GTR + F + I + I + R5$ that emerged as the best model through the model finder [6]. TreeTime version $0.82$ [7] was used to visualise the reconstructed national phylogenetic tree on a time scale while the trees for individual variants were visualised and coloured by region using an online tool for displaying, annotating and managing trees called an Interactive Tree Of Life (iTOL).

### 2 Results

#### 2.1 SARS-CoV-2 Epidmiology

##### 2.1.1 Cases

**Table 2.** Estimation results for the GAM model-cases, the intercept is significant

| | Estimate | Standard Error | $z-$value | $p-$value | CI |
|---|---|---|---|---|---|
| Intercept | $3.070814$ | $0.060623$ | $50.654$ | $< 2 \times 10^{-16}$ | $(3.01, 3.13)$ |
| DayOfWeek | $-0.001999$ | $0.013506$ | $-0.148$ | $0.882$ | $(-0.02, 0.01)$ |

Diagnostic test results for a model with $k = 45$ basis dimension: effective degrees of freedom, $= 35.9$ and $k-$index$= 0.94$ is close to $1$, a sign that the basis dimension is large enough. Diagnostic plots are shown in Figures 1a, 1b, 1c and 1d.

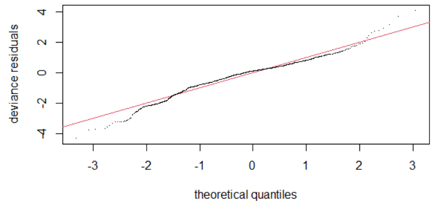


**(a)** Q-Q plot

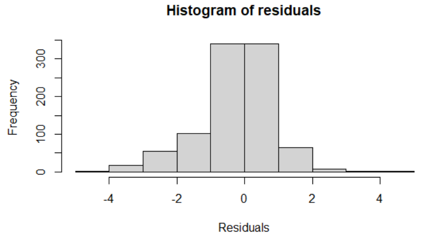


**(b)** Histogram

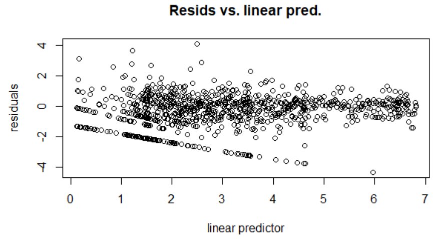


**(c)** Residuals versus Linear Predictor

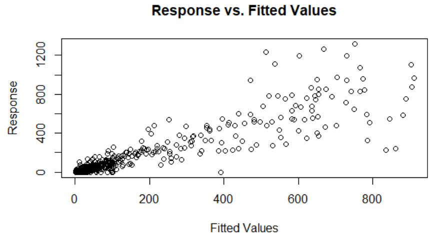


**(d)** Response against Fitted values

**Figure 1. Cases:** (a) Residuals almost follow a straight line. (b) A Bell-shaped histogram, indicating a good fit. (c) Points symmetric about $y = 0$. (d) Points are symmetric about $y = x$. All suggesting a good fit of the model

##### 2.1.2 Deaths

**Table 3.** Estimation results for the GAM model-deaths, intercept is significant

| | Estimate | Standard Error | $z-$value | $p-$value | CI |
|---|---|---|---|---|---|
| Intercept | $-0.50167$ | $0.09051$ | $-5.542$ | $2.98 \times 10^{-8}$ | $(-0.59, -0.41)$ |
| DayOfWeek | $0.01672$ | $0.01508$ | $1.109$ | $0.268$ | $(0.002, 0.03)$ |

Model diagnostic showed full convergence. There is a big gap between $k' = 43$ and the effective degrees of freedom, $edf = 30$ indicating sufficient basis dimension $k = 45$. $p - value = 1$ for day predictor is not significant, a sign that residuals are randomly distributed and there are enough basis functions. $k - index = 1.03$, as good as $1$, confirming a good choice for $k$. The model fits well as illustrated in diagnostic plots 2a, 2b, 2c and 2d.

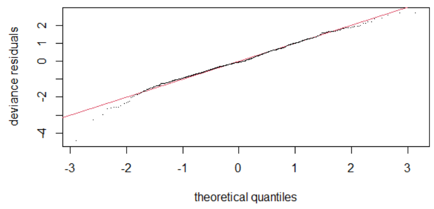


**(a)** Q-Q plot

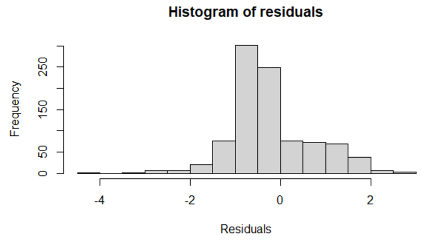


**(b)** Histogram

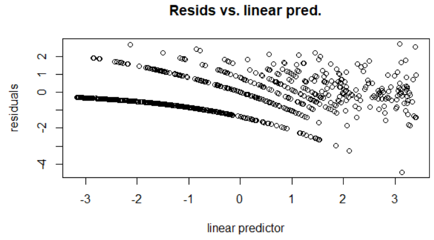


**(c)** Residuals against Linear Predictor

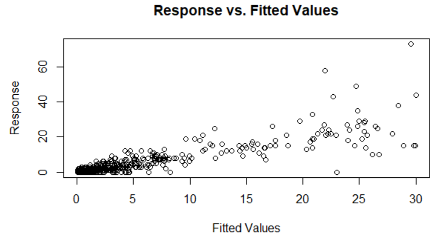


**(d)** Response versus Fitted values

**Figure 2. Deaths:** (a) Residuals follow a straight line. (b) The histogram is bell-shaped, indicating a good fit for the model. (c) Points are symmetric about $y = 0$. (d) Points are symmetric about $y = x$.

#### 2.2 Case fatality rate

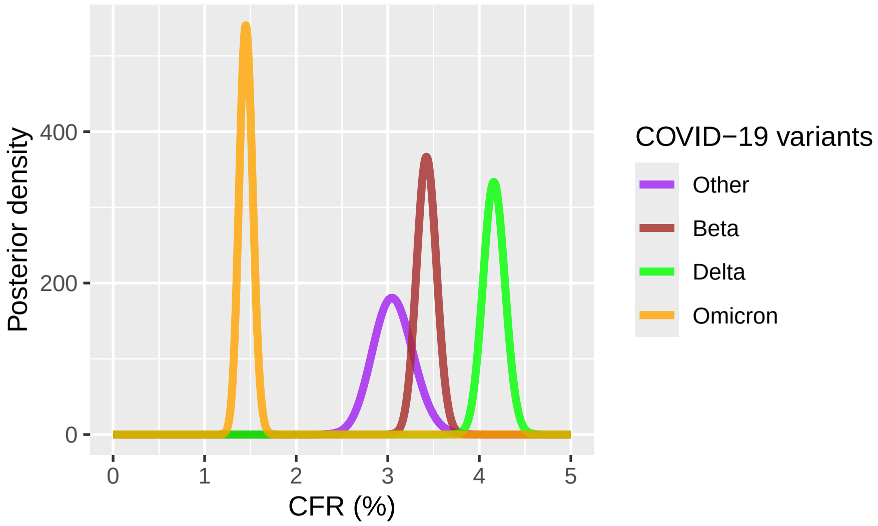


**Figure 3. Posterior distribution of CFR for the variants:** Omicron has the least CFR while Delta has the highest CFR.the distributions for Other and Beta variants almost overlap.

#### 2.3 Phylogenetic analysis

**Table 4.** Proportions and Confidence Interval of SARS-Cov-2 variants

| Variant | Southern region | | | Central region | | | Northern region | | | No region | | |
|---|---|---|---|---|---|---|---|---|---|---|---|---|
| | P | CI | SE | P | CI | SE | P | CI | SE | P | CI | SE |
| Alpha | 0.67 | | | 0 | | | 0 | | | 0.33 | | |
| Beta | 0.37 | 0.32, 0.41 | 0.05 | 0.16 | 0.13, 0.20 | 0.03 | 0.073 | 0.05, 0.10 | 0.02 | 0.40 | 0.34, 0.45 | 0.06 |
| Delta | 0.67 | 0.63, 0.71 | 0.04 | 0.07 | 0.05, 0.10 | 0.02 | 0.01 | 0.00, 0.02 | 0.01 | 0.24 | 0.21, 0.28 | 0.03 |
| Omicron | 0.78 | 0.73, 0.83 | 0.05 | 0.08 | 0.05, 0.12 | 0.03 | 0.14 | 0.10, 0.18 | 0.04 | 0.01 | 0.00, 0.03 | 0.01 |
| Other | 0.37 | 0.26, 0.48 | 0.11 | 0.04 | 0.01, 0.11 | 0.04 | 0.05 | 0.02, 0.13 | 0.04 | 0.55 | 0.44, 0.66 | 0.11 |

In Table 4, $P$ is the proportion, $CI$ is the $95\%$ confidence interval and $SE$ is the standard error.

#### 2.4 $\chi-$square test on SARS-CoV-2 proportions

**Table 5.** Distribution of SARS-Cov-2 variants across the regions

| Variant | Southern region | Central region | Northern region | No region | Total | Total (%) |
|---|---|---|---|---|---|---|
| Alpha | 4 | 0 | 0 | 2 | 6 | 0.42 |
| Beta | 180 | 78 | 36 | 198 | 492 | 34.6 |
| Delta | 383 | 42 | 5 | 139 | 569 | 40.01 |
| Omicron | 213 | 21 | 37 | 2 | 273 | 19.2 |
| Other | 25 | 3 | 2 | 45 | 82 | 5.77 |

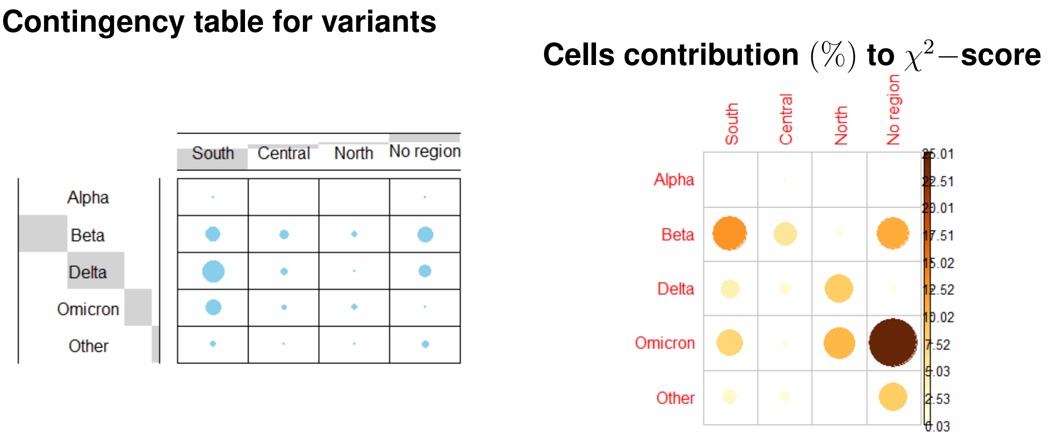


**(a)** Contingency table of COVID-19 variants

**(b)** cell contribution in percent to Chi-square test score

**Figure 4.** Contingency table and residuals plots
